# Fetal macrosomia and its associated factors among pregnant women delivered at national referral hospital in Uganda, a case-control study

**DOI:** 10.1101/2023.11.05.23298121

**Authors:** Peter Wanyera, Eve Nakabembe, Mike Nantamu Kagawa

**Affiliations:** Department of obstetrics and gynaecology, School of Medicine, College of Health Sciences, Makerere University. P.O. Box 7072, Kampala, Uganda; Busiu Health Centre IV, Mbale District Local Government, Uganda

**Keywords:** Fetal macrosomia, maternal weight, risk factors

## Abstract

**Background:** The delivery of macrosomic newborns (newborns>4000gm) is associated with many complications, yet the number macrosomic newborns is increasing steadily worldwide. Studies suggest the risk factors for fetal macrosomia include weight at first antenatal visit, previous delivery of a large newborn, newly diagnosed diabetes in pregnancy, increasing number of deliveries, a male fetus, and many others. The objective of this study was to determine the risk factors for fetal macrosomia among women who delivered at a National Referral Hospital in Kampala, Uganda in order to address a gap in knowledge in this area.

**Methods:** An unmatched case-control study was conducted among 177 cases and 354 controls at Kawempe National Referral Hospital. Data was collected using interviewer-administered questionnaires. Bivariate and multivariate analysis was done using STATA version 16.0.

**Results:** Risk factors for fetal macrosomia included maternal age ≥40 years (aOR = 7.4, [95%CI 1.37 - 39.44], p value = 0.020), maternal weight ≥80kg (aOR = 4.0, [95%CI 2.15 - 7.40], p value <0.001), maternal height ≥160cm (aOR = 1.6, [95%CI 1.02 - 2.51], p value = 0.040), being married (aOR = 2.55, [95%CI 1.08 - 6.06], P value = 0.038), gestation age ≥40 weeks (aOR = 1.8,[95%CI 1.16 – 2.82], p value = 0.009), previous macrosomia (aOR = 2.2, [95%CI 1.26 - 3.81], p value = 0.006) and male babies (aOR = 1.78, [95%CI 1.14 - 2.77], p value = 0.011)

**Conclusions:** Maternal demographic factors at the time of birth such as weight, height as well as advanced age significantly contribute to giving birth to large newborns. Other factors such as post-datism, previous delivery of a large newborn, male fetus and being in a marital relationship, were also noted. A well-designed protocol to identify women with risk factors for fetal macrosomia may help to provided targeted interventions in this group.

**Plain English Summary:** The delivery of large newborns (greater than 4000gm) is associated with many complications for both the mother and the newborn, and yet the number large newborns is increasing steadily worldwide.

Documented factors shown to increase the likelihood of delivering a large newborn include weight at first antenatal visit, previous delivery of a large newborn, increased weight gain during pregnancy, maternal obesity, newly diagnosed diabetes in pregnancy, pregnancies going beyond the due date, a male fetus, and advanced maternal age.

There is paucity of information regarding delivery of delivery of large newborns in Uganda. We therefore set out to determine the factors that increase the likelihood of delivering large newborns among women who delivered at the National Referral Hospital in Kampala, Uganda.

In this study we retrospectively compared 177 women with large newborns and 354 women who had average-sized newborns.

Our findings indicate an increased likelihood of delivering a large newborn among mothers who were greater than 80kg, more than 40 years and taller than 160cm, as well as those who were married, carrying a male infant, where the pregnancy went beyond 40 weeks, and those with a previous delivery of a large newborn.

Maternal demographic factors at the time of birth such as weight, height as well as advance in age could significantly contribute to giving birth to a large newborn. Other factors such as a pregnancy going beyond its due date, having previously delivered a large newborn, a male fetus and being in a marital relationship, were also noted.

## Introduction

The worldwide prevalence of birth of infants greater or equal to 4000gm is approximately 9 percent and 0.1 percent for weight greater or equal to 5000g, with wide variations among countries [1]. Globally, macrosomia affects 3 to 15% of all pregnancies [2] and in high income countries, the magnitude of macrosomia ranges from 5 to 20% of all births [3]. In the United States, approximately 7 percent of live born infants weigh greater than or equal to 4000 g and 1 percent weigh greater than 4500g [4]. The prevalence of birth weight greater than or equal to 4000 g in low-income countries is typically 1 to 5 percent but ranges from 0.5 to 14.9 percent [5]. For example, findings from a recent study showed a prevalence of macrosomia of 6.7% in Ethiopia [6].

Management of fetal macrosomia has for long been an obstetric challenge and is becoming an increasingly important problem because of the rising incidence of macrosomia and the associated risks to the mother and infant. Recent evidence suggests that the incidence of macrosomia is increasing. This is attributed to the increase in maternal anthropometry, reduced cigarette smoking, and changes in socio -demographic factors [7].

Documented risk factors for macrosomia are weight at first visit, previous macrosomic infant, excessive weight gain in pregnancy, obesity, gestational diabetes, multiparity, post-datism, male fetus, and advanced maternal age [8, 9]. Other factors associated with fetal macrosomia include genetics, duration of gestation, presence of gestational diabetes, and diabetes mellitus types I and II. Genetic, racial, and ethnic factors influence birth weight and the risk of macrosomia [10]. Fetal macrosomia complicates delivery process for both mothers and neonates and macrosomic babies have higher rates of developing both short- and long-term adverse health outcomes [2]. Fetal and neonatal outcomes of fetal macrosomia include shoulder dystocia, birth trauma like brachial plexus injury and skeletal injury, chorioamnionitis because of prolonged labour, meconium aspiration, low APGAR score, neonatal hypoglycemia and intrauterine fetal death (8).

Since prior diagnosis / antenatal prediction of fetal macrosomia is challenging both by sonography and usual estimation by palpation, paying attention to detail in terms of risk assessment and tests like blood sugar levels, maternal weight, height, previous history in terms of birth outcome and birth weights could help identify more of these mothers. [11]. The aim of this study was to determine the risk factors for fetal macrosomia among women who delivered at Kawempe National Referral Hospital (KNRH) in Kampala, Uganda. It is hoped that with this information, it will be possible to suggest ways in which prediction of macrosomia can be improved in order to institute interventions to minimise the adverse maternal and fetal complications associated with macrosomia.

## Materials and Methods

This was an unmatched case-control study conducted among postpartum women on the labour and post-natal wards at KNRH in Kampala. KNRH is one of the largest and public national referral hospitals in Uganda. It is a teaching hospital for the department of Obstetrics and Gynecology of Makerere University College of Health Sciences. The hospital is located approximately 12 kilometers from the capital city centre and has a bed capacity of 170. The population of Kampala City where the hospital is located is about 2 million people during the night, but this rises to about 4.5 million during the day. About 60-80 mothers are delivered daily at the hospital with 40-60 spontaneous vaginal deliveries and 15-20 caesarian deliveries with 2-3 macrosomic babies daily. Most patients received are from within the city and the surrounding districts as well as referrals from lower health units all over the country.

### Characteristics of participants

Participants were women who delivered at KNRH during the study period. Women with babies of birth weight greater or equal to 4000gm were taken as cases and those with babies weighing 2500kg – 3990kg were considered as controls.

### Sample Size Determination

The sample size was estimated using an online formula for case control studies by Glaziou Phillipe [12]. This was based on a study done in Ethiopia by Wondie et al to determine factors associated with macrosomia among neonates delivered at Debre Markos Referral Hospital, Northwest Ethiopia in 2014 [13]. In this study, 70.7% of multiparous mothers had macrosomic babies, compared to 60.1% of the controls. Using the following assumptions: Odds ratio 1.9, Exposed controls 60.1%, Alpha risk 5%, and a Control / Case ratio 2:1, the number of cases required was determined to be 177, and the number of controls 354; giving an overall sample size of 531 participants.

### Sampling Method

Consecutive sampling was used to recruit cases. A case was any mother in the postnatal ward with a baby birth weight of 4000gm or more as documented in the delivery register. For each case, two controls were selected and these included mothers who had delivered a baby weighing between 2500 – 3999gm, just before and after the case as indicated in the delivery register giving a ratio of cases: controls = 1:2. In case any of the selected controls as per the register was found to be a case, then four controls were selected, that is two before and two after the two cases.

During sampling, mothers who were identified as cases or controls were approached and given a full explanation about the study. Following acceptance to participate in the study, written informed consent was obtained. For minors (emancipated or non-emancipated), consent was sought from a legal guardian or parent present at time of recruitment. Data collection was done within 24hours of delivery using a pre-tested interviewer administered questionnaire. Additional Information was obtained from the mother’s clinical chart/delivery report where such information was documented.

### Data collection and analysis

Data was collected using pre-tested interviewer administered structured questionnaires for three months from 8^th^ February 2021 to 25^th^ May 2021 when the expected sample size was achieved. A database was designed using the computer software EPI-DATA version 4.6 for data. The raw data was securely stored to maintain confidentiality. The data was then exported to STATA version 16.0 software for analysis. For continuous data that was normally distributed, comparison between the cases and the controls was done using the student t-test. For data that was not normally distributed, comparison between the cases and the controls was done using the Mann Whitney U Test (Wilcoxon Rank Sum Test). Binary logistic regression was used in multivariate analysis for factors associated with fetal macrosomia. All variables with a P value <0.05 using the chi-square test were included in the multivariable model. Backward stepwise selection (or backward elimination) method was used in analysis for the associated factors. Odds ratios were used as the measure of association and reported along with their 95% confidence intervals.

### Ethics approval and consent to participate

approval was obtained from the Makerere University School of Medicine Research and Ethics Committee (#REC REF 2021-002). Voluntary written informed consent was sought from each participant before enrolment into the study. If the participant was a minor, the parent or legal guardian was requested to provide an informed consent in writing before the subject was enrolled into the study. The study participants were informed of their right to refuse participation or withdraw from the study at any time if they so wished, and that refusal to participate or withdrawal of consent from the study would not result in any penalties or have any impact on their care in the hospital.

### Consent for publication

the participants were informed during the consent process that the findings, without disclosing their names may be published, and they had no objection. All authors consent to be a part of this publication by taking full responsibility and accountability for the contents of the article. All the work, figures and tables in this publication were prepared by the authors and where other work is used, it has been appropriately cited.

## Results

A total of 531 participants were recruited into the study with 177 (33.3%) as cases and 354 (66.7%) as controls making up a ratio of 1:2. The maternal age, weight and height was significantly higher among the cases compared to the controls across all the categories (Table 1).

**Table 1.**
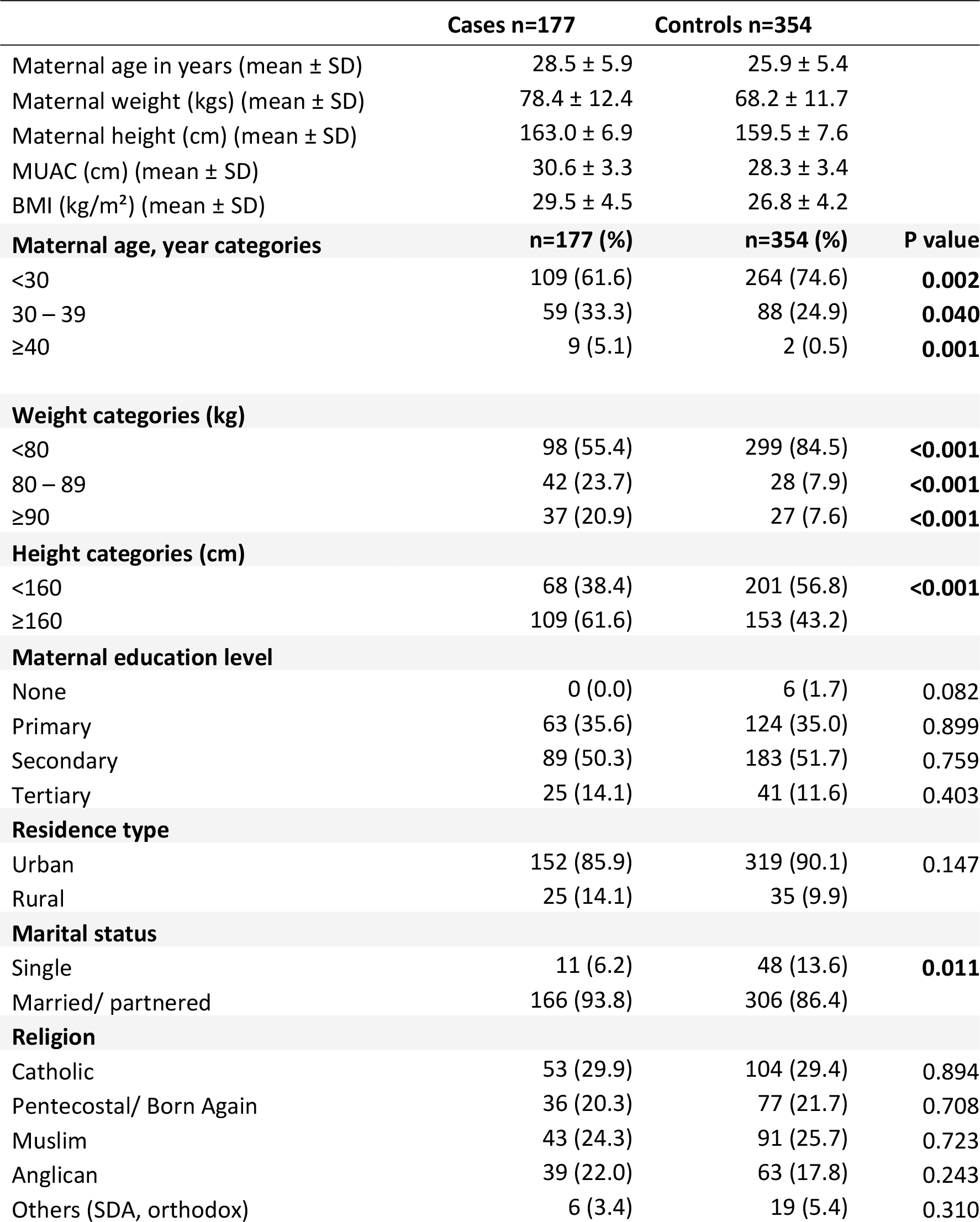
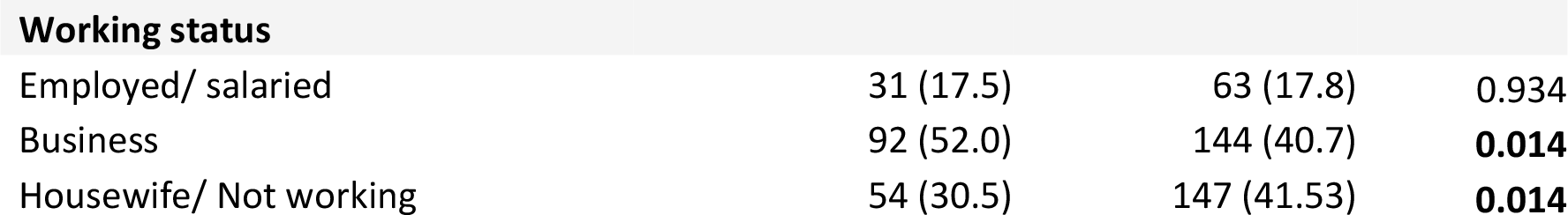
Maternal characteristics.

|  | **Cases n=177** | **Controls n=354** |  |
| --- | --- | --- | --- |
| Maternal age in years (mean ± SD) | 28.5 ± 5.9 | 25.9 ± 5.4 | |
| Maternal weight (kgs) (mean ± SD) | 78.4 ± 12.4 | 68.2 ± 11.7 | |
| Maternal height (cm) (mean ± SD) | 163.0 ± 6.9 | 159.5 ± 7.6 | |
| MUAC (cm) (mean ± SD) | 30.6 ± 3.3 | 28.3 ± 3.4 | |
| BMI (kg/m<sup>2</sup>) (mean ± SD) | 29.5 ± 4.5 | 26.8 ± 4.2 | |
| **Maternal age, year categories** | **n=177 (%)** | **n=354 (%)** | **P value** |
| <30 | 109 (61.6) | 264 (74.6) | **0.002** |
| 30 – 39 | 59 (33.3) | 88 (24.9) | **0.040** |
| ≥40 | 9 (5.1) | 2 (0.5) | **0.001** |
| **Weight categories (kg)** |  |  |  |
| <80 | 98 (55.4) | 299 (84.5) | **<0.001** |
| 80 – 89 | 42 (23.7) | 28 (7.9) | **<0.001** |
| ≥90 | 37 (20.9) | 27 (7.6) | **<0.001** |
| **Height categories (cm)** |  |  |  |
| <160 | 68 (38.4) | 201 (56.8) | **<0.001** |
| ≥160 | 109 (61.6) | 153 (43.2) | |
| **Maternal education level** |  |  |  |
| None | 0 (0.0) | 6 (1.7) | 0.082 |
| Primary | 63 (35.6) | 124 (35.0) | 0.899 |
| Secondary | 89 (50.3) | 183 (51.7) | 0.759 |
| Tertiary | 25 (14.1) | 41 (11.6) | 0.403 |
| **Residence type** |  |  |  |
| Urban | 152 (85.9) | 319 (90.1) | 0.147 |
| Rural | 25 (14.1) | 35 (9.9) |  |
| **Marital status** |  |  |  |
| Single | 11 (6.2) | 48 (13.6) | **0.011** |
| Married/ partnered | 166 (93.8) | 306 (86.4) |  |
| **Religion** |  |  |  |
| Catholic | 53 (29.9) | 104 (29.4) | 0.894 |
| Pentecostal/ Born Again | 36 (20.3) | 77 (21.7) | 0.708 |
| Muslim | 43 (24.3) | 91 (25.7) | 0.723 |
| Anglican | 39 (22.0) | 63 (17.8) | 0.243 |
| Others (SDA, orthodox) | 6 (3.4) | 19 (5.4) | 0.310 |
| **Working status** |  |  |  |
| Employed/ salaried | 31 (17.5) | 63 (17.8) | 0.934 |
| Business | 92 (52.0) | 144 (40.7) | **0.014** |
| Housewife/ Not working | 54 (30.5) | 147 (41.53) | **0.014** |

The following characteristics were significantly higher among the cases than in the controls; gestation age (39.5 ± 1.1 vs 39.0 ± 1.3, P value <0.001), ever delivered a baby greater than 4kg (30.5% vs 12.4%, P value <0.001) and use of herbal medicine during pregnancy (33.9% versus 21.7%, P value = 0.003) (Table 2).

**Table 2.** Pregnancy and birth related characteristics.

|  | **Cases**<br>**n=177 (%)** | **Controls**<br>**n=354 (%)** | **P value** |
| --- | --- | --- | --- |
| Gestation age in weeks (mean ± SD), n = 478 | 39.5 ± 1.1 | 39.0 ± 1.3 | **<0.001** |
| Gestation age categories (weeks) n = 478 |  |  |  |
| 37 – 39 | 83 (52.9) | 213 (66.4) | **0.004** |
| 40 – 42 | 74 (47.1) | 108 (33.6) |  |
| Parity (carried above 28 weeks) |  |  |  |
| 1 | 38 (21.5) | 123 (34.8) | **0.002** |
| 2 – 4 | 107 (60.4) | 192 (54.2) | 0.257 |
| $\geq 5$ | 32 (18.1) | 39 (11.0) | 0.054 |
| Ever delivered a baby >4kg |  |  |  |
| No | 123 (69.5) | 310 (87.6) | **<0.001** |
| Yes | 54 (30.5) | 44 (12.4) |  |
| **Number of deliveries >4kg (n=98)** |  |  |  |
| 1 | 36 (66.7) | 30 (68.2) | 0.874 |
| 2 | 13 (24.1) | 10 (22.7) | 0.876 |
| 3 | 4 (7.4) | 3 (6.8) | 0.910 |
| 4 | 1 (1.9) | 1 (2.3) | 0.884 |
| **Ever had a still birth** |  |  |  |
| No | 171 (96.6) | 345 (97.5) | 0.557 |
| Yes | 6 (3.4) | 9 (2.5) |  |
| **Number of still births (n=15)** |  |  |  |
| 1 | 4 (66.7) | 8 (88.9) | 0.143 |
| 2 | 2 (33.3) | 0 (0.0) |  |
| 3 | 0 (0.0) | 1 (11.1) |  |
| **Early neonatal death** |  |  |  |
| No | 176 (99.4) | 348 (98.3) | 0.281 |
| Yes | 1 (0.6) | 6 (1.7) |  |
| **Use of herbal medicine during pregnancy** |  |  |  |
| No | 117 (66.1) | 277 (78.3) | **0.003** |
| Yes | 60 (33.9) | 77 (21.7) |  |

The mean Apgar scores at 5 minutes was significantly lower for cases as compared to controls (8.4 ± 2.6 vs 9.0 ± 2.0, P value = 0.009). In addition, the proportion of male babies born to the cases was significantly higher than that for the controls (64.6% vs 55.3%, P value = 0.041) (Table 3).

**Table 3.** Delivery and fetal characteristics.

|  | **Cases**<br>**n=177 (%)** | **Controls**<br>**n=354 (%)** | **P value** |
| --- | --- | --- | --- |
| Neonatal Apgar at 1 minute (mean ± SD) | 7.2 ± 2.4 | 7.7 ± 1.9 | **0.004** |
| Neonatal Apgar at 5 minutes (mean ± SD) | 8.4 ± 2.6 | 9.0 ± 2.0 | **0.009** |
| **Mode of delivery** |  |  |  |
| Vaginal | 80 (45.2) | 159 (44.9) | 0.951 |
| Caesarean | 97 (54.8) | 195 (55.1) |  |
| **Labour complications** |  |  |  |
| None | 131 (74.0) | 264 (74.6) | 0.887 |
| Obstructed labor | 24 (13.6) | 39 (11.0) | 0.394 |
| Ruptured uterus | 7 (4.0) | 10 (2.8) | 0.485 |
| Others | 15 (8.5) | 41 (11.6) | 0.271 |
| **Labour augmentation with oxytocin** |  |  |  |
| No | 122 (70.9) | 252 (72.6) | 0.682 |
| Yes | 50 (29.1) | 95 (27.4) |  |
| **Current delivery outcomes** |  |  |  |
| Live birth | 164 (92.7) | 340 (96.1) | 0.094 |
| Still birth | 13 (7.3) | 14 (3.9) |  |
| **Sex of the baby** |  |  |  |
| Male | 113 (63.8) | 195 (55.1) | 0.054 |
| Female | 63 (35.6) | 159 (44.9) | **0.040** |
| Sexual development disorder | 1 (0.6) | 0 (0.0) |  |

### Risk factors for fetal macrosomia

On multivariate analysis, the following factors were found to be risk factors for fetal macrosomia; maternal age ≥ 40 years, maternal weight ≥ 80kg, maternal height ≥160cm, being married, gestation age ≥ 40 weeks, history of having a prior delivery of a baby greater than 4kg, use of herbal medicine during pregnancy, and having a male baby (Table 4).

**Table 4.** Multivariate analysis of maternal risk factors for fetal macrosomia.

|  | Cases<br>n (%) | Controls<br>n (%) | Adjusted OR<br>(95%CI) | P value |
| --- | --- | --- | --- | --- |
| **Number of participants** | 177 (33.3) | 354 (66.7) |  |  |
| **Maternal age, year categories** |  |  |  |  |
| <30 | 109 (61.6) | 264 (74.6) | Ref |  |
| 30 – 39 | 59 (33.3) | 88 (24.9) | 0.8 (0.49 - 1.38) | 0.456 |
| ≥40 | 9 (5.1) | 2 (0.5) | 7.4 (1.37 - 39.44) | 0.020 |
| **Weight, kg categories** |  |  |  |  |
| <80 | 98 (55.4) | 299 (84.5) | Ref |  |
| 80 – 89 | 42 (23.7) | 28 (7.9) | 4.0 (2.15 - 7.40) | <0.001 |
| ≥90 | 37 (20.9) | 27 (7.6) | 3.9 (1.99 - 7.49) | <0.001 |
| **Height, cm categories** |  |  |  |  |
| <160 | 68 (38.4) | 201 (56.8) | Ref |  |
| ≥160 | 109 (61.6) | 153 (43.2) | 1.6 (1.02 - 2.51) | 0.040 |
| **Marital status** |  |  |  |  |
| Single | 11 (6.2) | 48 (13.6) | Ref |  |
| Married/ partnered | 166 (93.8) | 306 (86.4) | 2.5 (1.05 - 5.95) | 0.038 |
| **Gestation age, weeks categories** |  |  |  |  |
| 37 – 39 | 83 (52.9) | 213 (66.4) | Ref |  |
| 40 – 42 | 74 (47.1) | 108 (33.6) | 1.8 (1.16 - 2.82) | 0.009 |
| **Ever delivered a baby >4kg** |  |  |  |  |
| No | 123 (69.5) | 310 (87.6) | Ref |  |
| Yes | 54 (30.5) | 44 (12.4) | 2.2 (1.26 - 3.81) | 0.006 |
| **Use of herbal medicine during pregnancy** |  |  |  |  |
| No | 117 (66.1) | 277 (78.3) | Ref |  |
| Yes | 60 (33.9) | 77 (21.7) | 2.3 (1.41 - 3.82) | 0.001 |
| **Sex of the baby** |  |  |  |  |
| Female | 63 (35.6) | 159 (44.9) | Ref |  |
| Male | 113 (63.8) | 195 (55.1) | 1.78 (1.14 - 2.77) | 0.011 |

## Discussion

This was an unmatched hospital-based case control study to identify the risk factors for fetal macrosomia among women delivering at a National Referral Hospital in Kampala, Uganda. Risk factors for fetal macrosomia were- maternal age greater or equal to 40 years, maternal weight of 80kg and above, height greater than 160cm, being married, gestational age greater or equal to 40 weeks, history of previous macrosomic baby, use of herbal medicines during pregnancy and a male fetus.

The study found out that maternal age of 40 years and above had increased odds of fetal macrosomia. Mothers who were aged 40 years and above were 7.4 times more likely to deliver a macrosomic baby compared to those below 40 years (aOR = 7.4, 95%CI 1.37 - 39.44, P value = 0.020). These findings are consistent with a study done in Nigeria where the mean age of mothers that had macrosomic babies was reported to be significantly higher than that of mothers with normal birth weight babies [14]. This could be related to the frequent development of medical conditions like Gestational diabetes mellitus in older women. This was however in disagreement with findings of a study done in South Africa which found that advanced maternal age women was associated with delivery of low birth weight babies [15].

Compared to women with maternal weight of 80kg or less, women with a maternal weight of 80-89kg had 4 times higher odds of delivering a macrosomic newborn (aOR = 4.0, 95%CI 2.15 - 7.40, P value <0.001),. This could be attributed to excess fats in obese mothers being broken down into glucose which is later transferred to the fetus across the placenta [16]. The findings from this study are similar to findings from studies done in Tanzania and Southeast Nigeria which reported an increased risk of macrosomia with increasing maternal weight [17, 18].

Maternal height greater or equal to 160cm was an independent risk factor for fetal macrosomia. Mothers whose height was greater or equal to 160cm were 60 percent more likely to deliver macrosomic babies compared to mothers whose height was less than 160cm (aOR = 1.6, 95%CI 1.02 - 2.51, P value = 0.040). These findings are similar to studies done in Nigeria and Saudi Arabia where it was reported that maternal height greater than 160cm was significantly associated with macrosomia compared to controls [14, 19, 20].

The odds of having a macrosomic infant were 2.5 times among the married women than in single women (aOR = 2.5, 95%CI 1.05 - 5.95, P value = 0.038). These findings are similar to results of studies done in Ethiopia and other low-and middle-income countries [6, 21]. This may be attributed to better social and economic support to aid in nutrition and psychological wellbeing of the mother and generally good quality of life [22].

In this study, mothers whose gestational age was greater or equal to 40 weeks were 1.8 times more likely to deliver a macrosomic baby compared to those less than 40 weeks of gestation (aOR = 1.8, 95%CI 1.16 – 2.82, P value = 0.009). This is similar to findings by Koyanagi et al in his study entitled macrosomia in 23 developing countries: an analysis of a multi-country, facility-based, cross-sectional survey [5]. Adugna also found similar trends in his study done in Gondar, Ethiopia where mothers having a gestational age greater or equal to 40 weeks were 4.1 times more likely to be deliver macrosomic babies than mothers with less than 40 weeks [3]. This may be a consequence of continued supply of nutrients and oxygen-rich blood to the developing fetus beyond 40 weeks.

A previous history of delivering a macrosomic infant was found to be associated with 2.2 times odds of having another macrosomic infant compared to mothers with no prior history (aOR = 2.2, 95%CI 1.26 - 3.81, P value = 0.006). This finding has been consistent across several studies [3, 23, 24]. This may be due to an inherited genetic tendency of a mother to deliver macrosomic babies. Use of herbal medicines during pregnancy was also found to be associated with macrosomia with a 2.3 times risk compared to mothers who never used herbal medicines herbs (aOR = 2.3, 95%CI 1.41 - 3.82, P value = 0.001). This is in contrast to findings from other studies that did not report any association [25, 26]. There seems no biologically plausible explanation except that these herbs may contain insulin inducing agents that may interfere with glucose metabolism hence leading to fetal macrosomia.

Delivery of a male infant was associated with macrosomia. Male babies were nearly 80 percent more likely to be macrosomic compared to their female counterparts (aOR = 1.78, 95%CI 1.14 - 2.77, P value = 0.011). These findings are consistent with other studies done in Africa which found that male gender was significantly associated with increased risk of macrosomia [27, 28]. Daily fetal growth appears to be higher in male fetuses than in females [29]. More weight gained by male babies in utero is thought to be a result of androgen action [30].

## Conclusions and recommendations

The risk factors for fetal macrosomia among women delivering at Kawempe National Referral Hospital in central Uganda were; maternal age greater or equal to 40 years, maternal weight of 80kg and above, height greater than 160cm, gestational age greater or equal to 40 weeks, history of previous macrosomic baby, being married, use of herbal medicines during pregnancy and male fetus. It therefore becomes imperative to identify pregnant women at risk of fetal macrosomia during ANC, admission, labour, using simple processes such measuring maternal weight and height so that effective education, precautions and interventions are implemented. A well-designed protocol to guide in the management of mothers at risk of macrosomia would go a long in reducing the adverse maternal and fetal outcomes such as shoulder dystocia, perineal trauma, fetal distress and many others that are known to be associated with fetal macrosomia.

### Study limitations

Pregnancy weight gain could not be established since most mothers did not know their pre-pregnancy weight.

## Data Availability

All data regarding this manuscript is available from the corresponding author upon reasonable request. This report is part of a thesis and data can be accessed with permission from the University.

## Declarations

### Availability of data and materials

All data regarding this manuscript is available from the corresponding author upon reasonable request.

### Authors’ contributions

All authors; WP, EN & MNK made significant contributions to this work from the conception stage, study design, execution, acquisition of data, analysis, and interpretation. MNK & WP drafted the initial manuscript and all authors participated in critically reviewing the article and made suggestions for improvement. All authors read and approved the final manuscript.

## Acknowledgements

We would like to acknowledge the support obtained from the management and staff of Kawempe National Referral Hospital, the work well done by the research assistants and all the study participants.

